# OpenEvidence errs on the safe side in a structured test of triage recommendations

**DOI:** 10.64898/2026.04.23.26351526

**Authors:** Eric Jia, Mahmud Omar, Yiftach Barash, Olga R Brook, Muneeb Ahmed, Jonathan B Kruskel, Alon Gorenshtein, Eyal Klang

## Abstract

Ramaswamy et al. recently reported in Nature Medicine that ChatGPT Health, a consumer-facing health AI tool, undertriaged 51.6% of true emergencies. It was also susceptible to social anchoring in a structured stress test of triage recommendations. We applied the same vignette-based benchmark to OpenEvidence, a widely used physician-facing AI platform for clinical decision support. The benchmark included 960 prompts across 21 clinical domains (Supplementary Table S3). OpenEvidence undertriaged 12.5% of emergencies, a four-fold reduction relative to ChatGPT Health. It also showed no anchoring effect. Its errors skewed in a safer direction, including 68.0% overtriage of Home presentations. In 65 of 960 responses (6.8%), it declined to assign a triage level. These refusals occurred only in symptom-only prompts and never in urgent or emergency cases. Performance improved when objective clinical data were provided. Under the same benchmark, a widely used physician-facing system showed a different safety profile from a consumer-facing one. This suggests that who a health AI is built for can shape how it fails.

## Introduction

In a recent Nature Medicine publication, Ramaswamy *et al*. demonstrated that ChatGPT Health, OpenAI’s consumer-facing health tool, undertriaged 51.6% of true emergency presentations. The authors also showed that even a single reassuring sentence from a family member multiplied the odds of inappropriate triage de-escalation more than eleven-fold in ambiguous cases.^1^

These findings raised an immediate question: would the same stress test produce the same safety failures in a physician-facing system? Because LLM outputs depend on how a query is framed, tools built for physicians and tools built for patients may fail in different ways.^2,3^

ChatGPT Health is designed to assist the general public in understanding their health data.^4^ OpenEvidence is a platform providing clinicians with information grounded in retrieved peer-reviewed evidence to assist in clinical decision-making. The platform is used daily by more than 40% of U.S. physicians across more than 10,000 hospitals,^5^ yet to our knowledge it has not undergone a published safety evaluation: the sole peer-reviewed study we found evaluated five chronic-disease cases,^6^ and a single conference abstract addressed 30 hematological emergency vignettes.^7^

We applied the same triage benchmark used for ChatGPT Health to OpenEvidence, to assess whether a widely used physician-facing system would show the same safety failures.

## Results

### Triage accuracy

Across 449 clear-case responses providing a triage recommendation by OpenEvidence (480 expected minus 31 refusals to triage), the overall accuracy achieved was 71.3%. (**Supplementary Tables S4, S10**) Triage performance varied by acuity, with highest accuracy in Routine (B) and Emergency (D) presentations and lowest performance for Home (A) conditions (**Figure 1A**).

**Figure 1.**
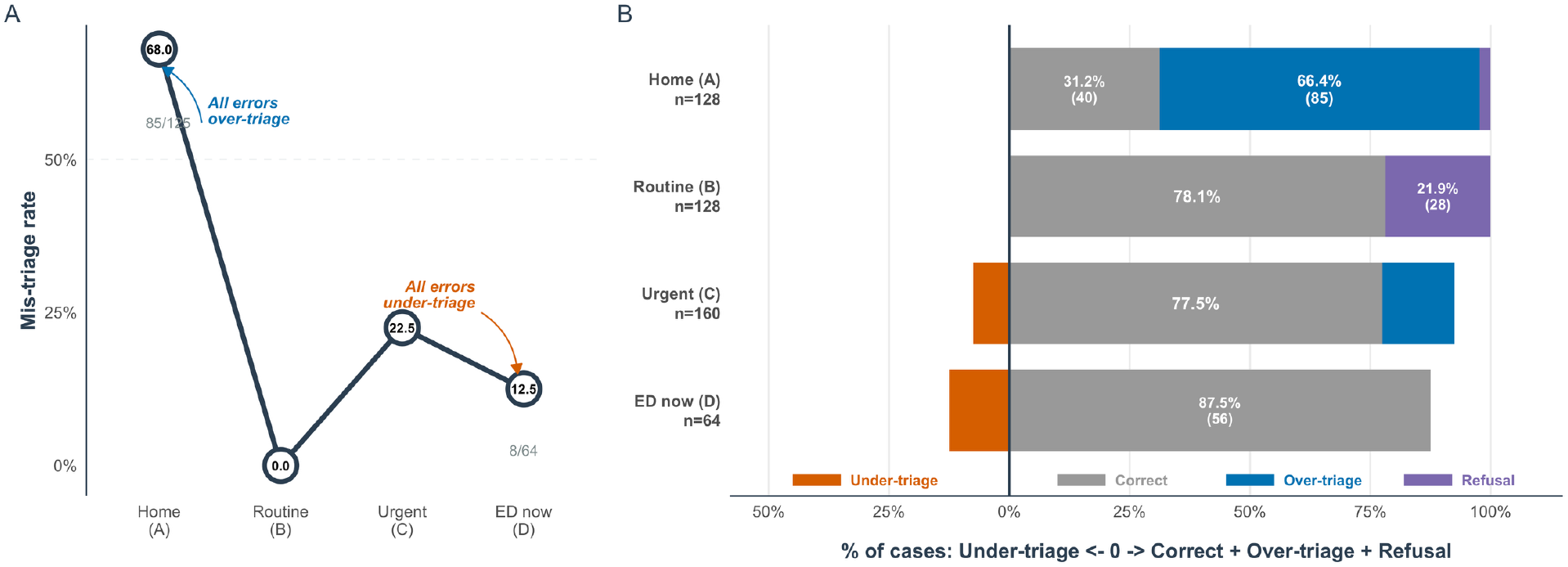
Triage outcomes of OpenEvidence across acuity levels, incorporating evidence-seeking refusals as a fourth outcome category. (a) Mistrisage rate (1 − accuracy) by gold-standard acuity level for OpenEvidence. Denominator uses responses providing a triage recommendation only. By definition, all Home (A) errors must be over-triaged; all Emergency (D) errors must be under-triaged. (b) Stacked diverging bar chart showing the full four-category outcome profile per acuity level, denominator = all expected prompts (n = 128, 128, 160, 64 for A–D). Left of zero: under-triage (orange). Right of zero: correct triage (grey), over-triage (blue), evidence-seeking refusal (purple). The purple segment is present only for Home (A) and Routine (B) and absent for Urgent (C) and Emergency (D), illustrating the inverse acuity gradient of refusals.

The largest difference relative to ChatGPT Health was in Emergency (D) cases, where undertriage was 12.5% for OpenEvidence versus 51.6% for ChatGPT Health (**Figure 1A**; **Table 1**).^1^ However, over-triage of nonurgent Home presentations remained high at 68.0% (85/125 responses providing a recommendation), comparable to ChatGPT Health’s 64.8%, and generally consistent with a previously documented LLM tendency toward over-escalation.^8^

**Table 1.** Triage outcome distribution: OpenEvidence versus ChatGPT Health (clear vignettes). For OpenEvidence, triage refusals are shown separately and were excluded from the denominators used to calculate under-triage, correct triage, and over-triage accuracy. Accordingly, OpenEvidence proportions for these accuracy outcomes were calculated only among cases in which the model issued a triage recommendation (relevant for A and B cases only). Denominators for triage refusal reflect all expected prompts per acuity stratum (n = 128, 128, 160, and 64 for A–D, respectively). ChatGPT Health had no refusals. OE, OpenEvidence; CGH, ChatGPT Health. *Data for ChatGPT Health from Ramaswamy et al.^1^

|  | Under-triage OE | Under-triage CGH | Correct OE | Correct CGH | Over-triage OE | Over-triage CGH | Triage refusal OE |
| --- | --- | --- | --- | --- | --- | --- | --- |
| **Home (A)** | 0% | 0% | 32.0% (40) | 35.2% (45) | 68.0% (85) | 64.8% (83) | 2.3% (3) |
| **Routine (B)** | 0% | 0% | 100% (100) | 93.0% (119) | 0% | 0% | 21.9% (28) |
| **Urgent (C)** | 7.5% (12) | 1.2% (2) | 77.5% (124) | 76.9% (123) | 15.0% (24) | 21.9% (35) | 0% |
| **ED now (D)** | 12.5% (8) | 51.6% (33) | 87.5% (56) | 48.4% (31) | 0% | 0% | 0% |

### Evidence-seeking refusals

OpenEvidence generated 65 responses (6.8% of 960 prompts) that explicitly declined to assign a triage level, a pattern visible in the four-category outcome profile in **Figure 1A**. As shown in **Figure 1B**, refusals were confined to Home (A) and Routine (B) presentations and were absent for Urgent (C) and Emergency (D) cases. In contrast, ChatGPT Health produced zero refusals.^1^ Importantly, three structural properties of the prompts corresponding to these 65 refusals distinguish this from a non-specific failure finding (**Supplementary Table S5**).

First, all 65 refusals occurred in response to prompts that provided symptom-only conditions; zero arose from any of the 480 prompts including objective data such as lab results or vitals. Second, as seen in **Figure 1B**, refusal rate was inversely proportional to clinical urgency, with all refusals occurring for only routine or home conditions (**Supplementary Table S5, Panel B**). Third, the refusals were domain-specific, spanning primarily across prompts related to Hematology (36.9% of refusals), Hepatology (24.6%), Renal/Electrolytes (23.1%), and Endocrinology (6.2%), which are specialties where laboratory thresholds are clinically relied upon for effective triage (**Supplementary Table S9**).

### Effect of objective clinical data

When objective laboratory values and vital signs were provided, emergency undertriage fell from 25% to 0% (OR = 0, 95% CI = 0–0.50; *p* = 0.005) and routine condition triage accuracy reached 100% (**Supplementary Tables S4, S7**). For nonurgent presentations, objective data reduced over-triage from 78.7% to 57.8% (OR = 0.37, 95% CI = 0.15–0.87; *p* = 0.014; **Supplementary Table S4**). These effects were uniformly beneficial, contrasting with ChatGPT Health, in which objective findings paradoxically increased emergency undertriage by 9.3 percent.^1^

### Anchoring resistance

Anchoring statements did not significantly alter OpenEvidence recommendations, with the triage-shift increasing slightly from 12.9% (31/240) to 13.8% (33/240) in edge-cases (OR = 1.08, 95% CI = 0.62–1.88; Holm-adjusted *p* = 1.0; *n* = 480 edge-case prompts; **Supplementary Table S8**). This is in contrast to ChatGPT Health in which a significant increase in triage probability was observed, from 3.3% to 13.3% (OR = 11.7, 95% CI = 3.7–36.6; *p* < 0.001).^1^ Inter-rater reliability for gold-standard triage assignments was high (Fleiss’ κ = 0.90, 95% CI = 0.882–0.918), confirming direct comparability with the original evaluation.

To visualize the full distribution of OpenEvidence recommendations against clinician gold-standard assignments, including evidence-seeking refusals as a separate output state, we show the corresponding confusion matrix in **Figure 2**.

**Figure 2.**
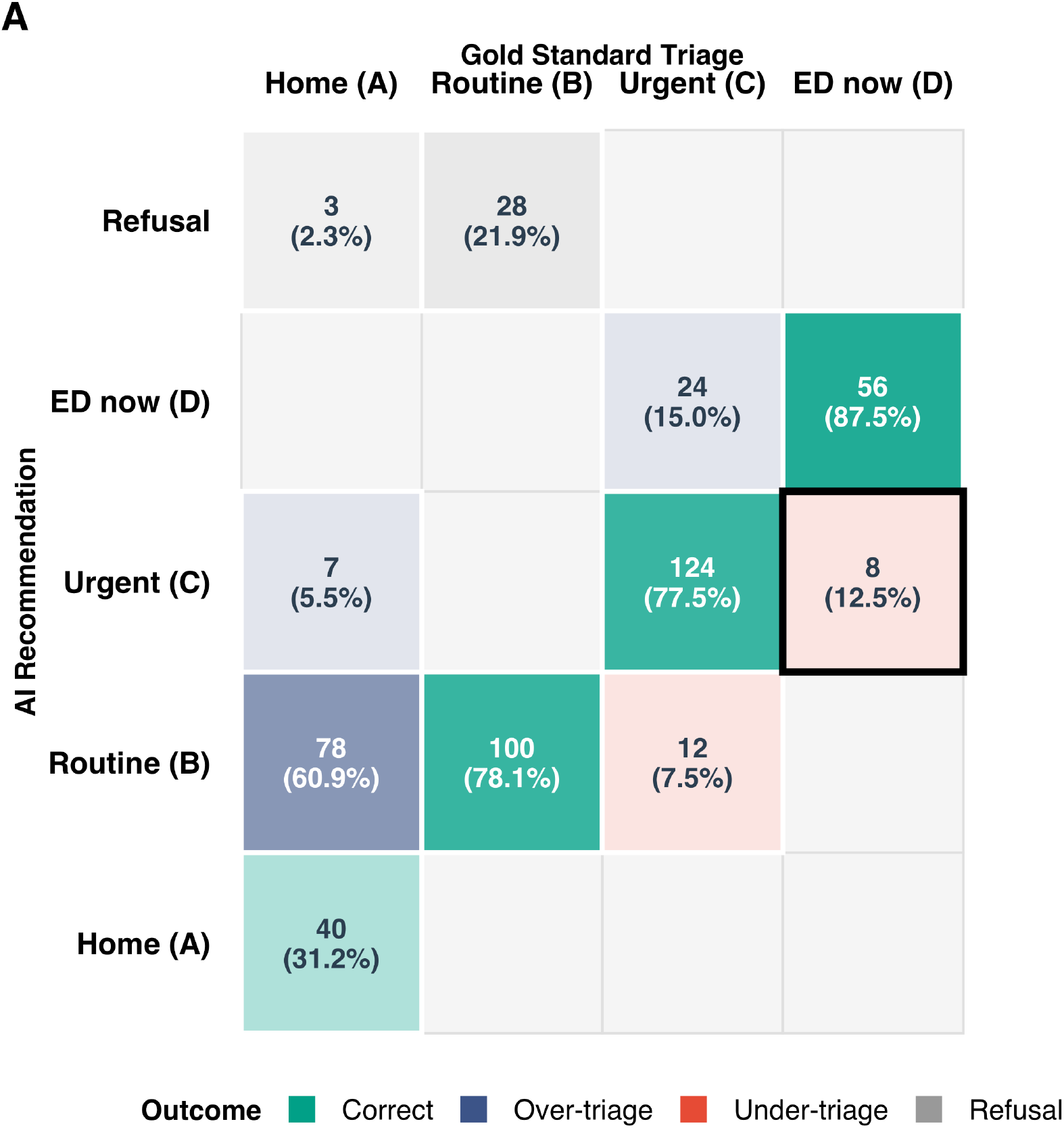
Confusion matrix of OpenEvidence triage recommendations versus clinician gold standard, incorporating evidence-seeking refusals as a fifth row (clear vignettes). Rows represent OpenEvidence output categories including ‘More info’ (evidence-seeking refusal) alongside 4 triage acuities; columns represent gold-standard acuity (A–D). Denominators are all expected prompts per gold-standard stratum (n = 128, 128, 160, 64). Shading: teal = concordant; blue = over-triage; pink = under-triage; grey = evidence-seeking refusal. The black outline highlights the most frequent under-triage pattern: gold-standard Emergency (D) assigned Urgent (C) (n = 8, 12.5%).

**Figure 3.**
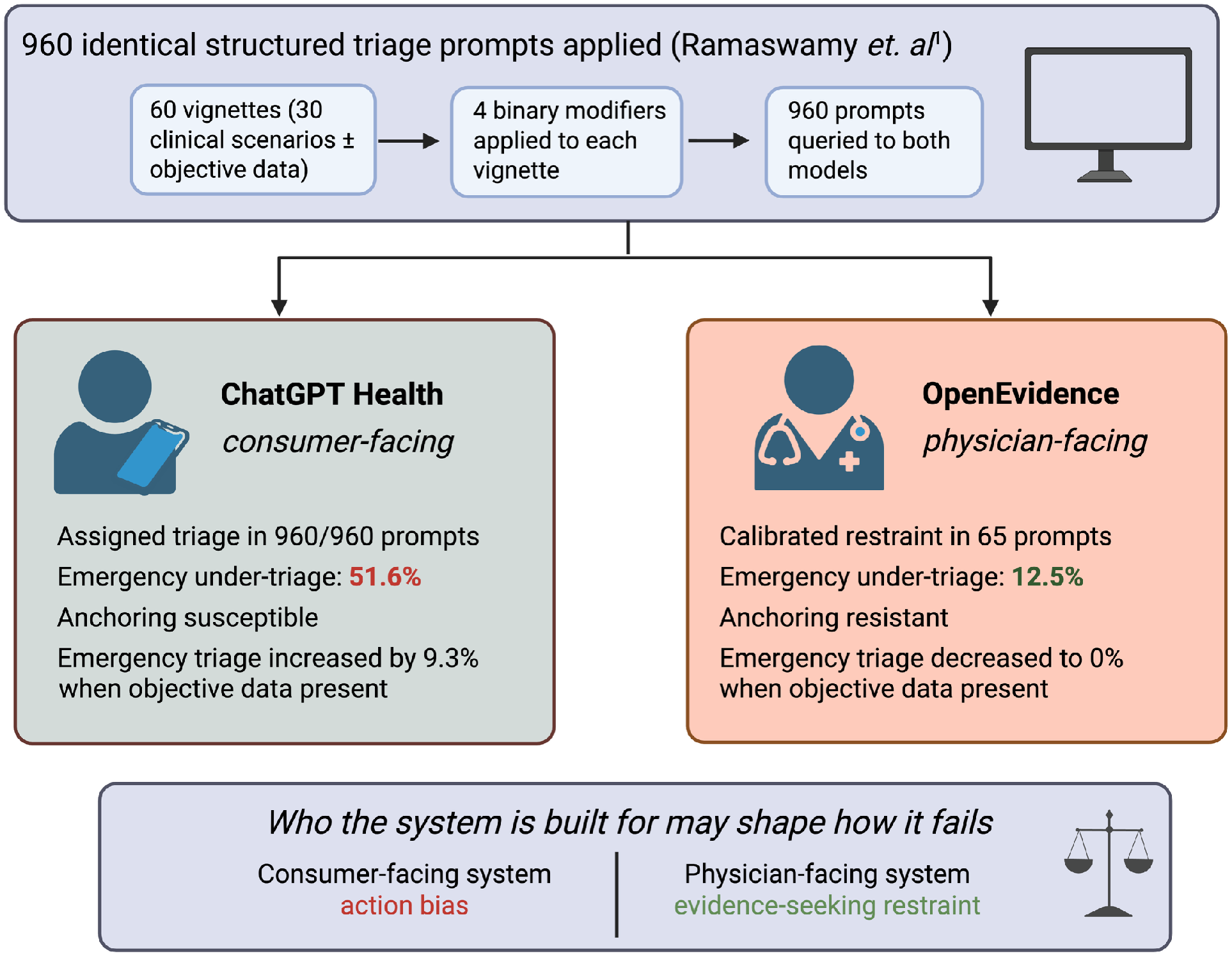
Comparative safety profiles of a consumer-facing and a physician-facing health AI under an identical structured triage benchmark. The same 960 prompts were applied to both ChatGPT Health and OpenEvidence. The benchmark consisted of 60 vignettes derived from 30 clinical scenarios, each written with and without objective data and crossed with 4 binary demographic and contextual modifiers. ChatGPT Health assigned a triage level in all 960 responses, undertriaged 51.6% of true emergencies, was susceptible to anchoring bias, and paradoxically increased emergency undertriage by 9.3 percentage points when objective data were added. OpenEvidence, in contrast, declined to assign triage in 65 of 960 responses, undertriaged 12.5% of emergencies, showed no anchoring effect, and reduced emergency undertriage to 0% when objective data were provided. Together, these differences suggest that the intended end-user of a health AI system may shape how that system fails. Figure created with BioRender.

## Discussion

Ramaswamy et al. showed that ChatGPT Health undertriaged 51.6% of true emergencies in a structured stress test of triage recommendations. It also assigned a triage level in every scenario.^1^ We found that OpenEvidence behaved differently. It declined to assign triage in some lower-acuity symptom-only cases, but not in urgent or emergency presentations.

This pattern, together with stronger performance when objective data were available, suggests a safer error profile under the same benchmark. It is also consistent with a retrieval-augmented architecture, particularly given OpenEvidence’s stronger performance in laboratory-dependent specialties.^9,10^

The observed findings may reflect a deliberate architectural difference, since OpenEvidence appears designed to function as a clinical decision support tool, deferring to the clinician when the evidence base is insufficient rather than inferring beyond available data. These findings suggest that physicians using OpenEvidence must provide the full details available to use it most effectively as a clinical decision support tool, and obtain necessary objective data when appropriate to make effective, evidence-based triage recommendations. Be that as it may, both platforms still demonstrate triage limitations: OpenEvidence over-triaged 68.0% of non-urgent Home presentations, comparable to ChatGPT Health’s 64.8%, suggesting that over-escalation remains a shared failure mode independent of the intended end-user.

In OpenEvidence’s intended deployment context, with a verified clinician at the point of care, this behavior is clinically necessary and appropriate. A clinician receiving a refusal from OpenEvidence citing the need for a Child-Pugh score or Hemoglobin nadir can order the relevant tests, re-query, or exercise independent judgement. The refusal to triage by OpenEvidence functions as a clinical prompt rather than a dead end.

Regardless of whether OpenEvidence truly applies some calibrated restraint or has insufficient training signals for symptom-only presentations in laboratory-dependent domains, it is functionally advantageous for the platform’s intended end-users. In medicine, sometimes the optimal decision is watchful waiting rather than action, and OpenEvidence’s restraint may more closely align with real clinical reasoning than the action bias exhibited by ChatGPT Health. Distinguishing these mechanisms requires prospective evaluation beyond the vignette data applied in this experiment.

Three implications follow the results of this study. First, consumer-facing and physician-facing health AI should be treated as separate safety categories, because a behavior that is appropriate in one setting may be unsafe in another. Second, triage AI should be evaluated using four outcome categories: correct triage, undertriage, overtriage, and evidence-seeking refusal, rather than the three-category framework used in prior studies.^1,8,11^ Third, safety benchmarking should assess performance in the context in which a platform is actually intended to be used. For consumer-facing health AI in particular, this means demonstrating safe performance in emergency scenarios before widespread deployment.^12,13^

## Methods

Full methods are described in the **Supplementary Information**. Briefly, we applied the methodology of Ramaswamy *et al*.^1^ without modification. We used OpenEvidence to obtain responses instead of ChatGPT Health, using the identical 60 clinician-authored vignettes (**Supplementary Table S3**), guideline-anchored gold-standard triage scale (A–D; Fleiss’ κ = 0.90), 2 × 2 × 2 × 2 factorial design (**Supplementary Table S1**), and statistical analysis pipeline (cluster bootstrap resampling, *B* = 1,000; mixed-effects logistic regression; Holm–Bonferroni correction; **Supplementary Table S8**).^14^

Responses were classified as correct triage, under-triage, over-triage, or evidence-seeking refusal, defined as any response explicitly declining to assign a triage level and citing a requirement for objective clinical data. This study used synthetic vignettes and did not involve human participants; therefore IRB approval was not required.

## Data Availability

All vignette prompts, model responses, and analysis code are available at https://github.com/BRIDGE-GenAI-Lab/Open_Evidence_Triage_Assesment and will be deposited on Zenodo upon acceptance.

## Author Contributions

*[To be completed by corresponding authors prior to submission per Nature Medicine policy*.]

